# Association between Gallstones or Cholecystectomy and Upper Gastrointestinal Cancers: A Systematic Review and Meta-analysis

**DOI:** 10.64898/2026.01.07.26343554

**Authors:** Fangqing Li, Gwen Murphy, Gemma Mortell, Amanda Cross, Rhea Harewood

**Affiliations:** Cancer Screening and Prevention Research Group, Department of Surgery and Cancer, Imperial College London, London, United Kingdom

**Keywords:** meta-analysis, gastric cancer, oesophageal cancer, gallstones, cholecystectomy, cancer prevention

## Abstract

**Background:** Gallstones and cholecystectomy may be associated with upper gastrointestinal cancer risk but findings from individual studies are inconsistent. We undertook a systematic review and meta-analysis of available evidence.

**Methods:** PubMed/MEDLINE and Embase were searched through September 2024 to identify case-control and cohort studies investigating gallstones or cholecystectomy and oesophageal or gastric cancer risk. Random effects models estimated pooled relative risks (RRs) and 95% confidence intervals (CIs) for oesophageal and gastric cancer. Heterogeneity was evaluated using I^2^ and τ^2^ statistics.

**Results:** This meta-analysis included 15 studies. There was no association between gallstones [RR (95%CI) = 1.05 (0.97-1.14)] or cholecystectomy [RR (95% CI) = 0.99 (0.87-1.13)] and oesophageal cancer; however, both were positively associated with gastric cancer [RR (95% CI) = 1.25 (1.08-1.44); 1.10 (1.05-1.16), respectively]. In subgroup analyses, cholecystectomy was positively associated with non-cardia [RR (95%CI) = 1.17 (1.04-1.33)] but not cardia gastric cancer [RR (95% CI) = 0.89 (0.78-1.02)]; there were no other statistically significant associations by subtype/subsite, however data were limited for some subgroups.

**Conclusions:** Gallstones and cholecystectomy were positively associated with gastric cancer, cholecystectomy was specifically associated with non-cardia gastric cancer. Endoscopic screening for gastric cancer may be considered among individuals with gallstones or cholecystectomy history. PROSPERO registration: CRD42024563020

## Background

Oesophageal and gastric cancer are leading causes of cancer incidence and mortality worldwide [1, 2]. These upper gastrointestinal cancers are highly fatal diseases, with an overall 5-year survival of less than 20% [3]. Early-stage diagnosis of these malignancies is difficult due to the late onset of symptoms [2]. Consequently, most of these tumours are diagnosed at an advanced stage when treatment options are limited, and curative treatment is almost absent, leading to poor survival [2]. Identifying risk factors for these malignancies to inform primary and secondary prevention efforts is crucial.

Upper gastrointestinal cancer subtypes have distinct aetiologies. Oesophageal cancer is categorised into two primary histologic subtypes: adenocarcinoma (OAC) and squamous cell carcinoma (OSCC). Gastric cancer is commonly divided into two topographical subsites: gastric cardia (GCC) and non-cardia cancer (NCGC). These subtypes have distinct risk factors [1], though OAC and GCC share a common risk factor of chronic gastro-oesophageal reflux disease (GORD) [4]. Experimental studies have shown that bile contents in duodenogastric and gastroesophageal refluxate play a key role in GORD-associated carcinogenesis [5]. Therefore, inflammatory conditions that disturb bile regulation might increase the risk of these cancers, and the risks might differ by cancer subtype.

Gallstones refer to pebble-like concretions formed in the biliary tract and are one of the most prevalent digestive disorders worldwide [6]. Cholecystectomy is the standard treatment for symptomatic gallstones [7], and approximately 22% of cholecystectomy procedures are performed for gallstones [8]. Gallstones and cholecystectomy can disrupt physiological processes of the digestive system and have been linked to various chronic diseases including diabetes [9], cardiovascular diseases [10], and gastrointestinal cancers [11].

Previous studies have investigated gallstones or a history of cholecystectomy and the risk of upper gastrointestinal cancers, with mixed findings. Three systematic reviews and meta-analyses have been conducted on cholecystectomy and oesophageal or gastric cancer [12–14]; however, none of these differentiated between cholecystectomy performed for gallstones and those conducted for other gallbladder diseases. This suggests that the observed positive associations for cholecystectomy may be confounded by the effects of concurrent gallstones, if an effect of gallstones on the risk of upper gastrointestinal cancers indeed exists. To our knowledge, there have been no meta-analyses on cholecystectomy and oesophageal cancer since 2012 [13] and there are no systematic reviews or meta-analyses on gallstones and either oesophageal or gastric cancers. We investigated the association between gallstones, cholecystectomy or a combined exposure with upper gastrointestinal cancer risk by subtype and subsite.

## Methods

### Exposures of interest

We reviewed three categories of exposure: gallstones, cholecystectomy, and a combined exposure of both. Gallstones refers to reports of screening-detected or symptomatic gallstones in patients who had not undergone a cholecystectomy, as well as gallstones cases from studies where the cholecystectomy status was not reported. Cholecystectomy includes both open and laparoscopic cholecystectomy performed for all benign gallbladder diseases. Cases where cholecystectomy was specifically conducted as a treatment for gallstones were included in the combined exposure group.

### Search strategy and data collection

A systematic literature search was conducted in two major databases (PubMed/MEDLINE and Embase) from inception to September 2024 with the following combination of search terms: (cholelithiasis OR gallstone OR choledocholithiasis OR cholecystolithiasis OR cholecystectomy) AND (oesophageal OR stomach OR gastric) AND (cancer OR carcinoma OR tumour OR neoplasm OR adenocarcinoma). Medical subject headings, wildcards and proximity searches were used to increase search coverage. The British Medical Journal Best Practice search filters for case-control studies and cohort studies were incorporated into both database searches [15]. Additionally, citation tracking was performed to identify additional eligible studies by manually screening the reference lists of the included studies and previous systematic reviews.

Inclusion criteria were: (1) original studies using cohort or case-control study designs; (2) an outcome of incident oesophageal or gastric cancer; (3) adult participants (aged ≥ 18 years); (4) a patient population diagnosed with gallstones or with a history of cholecystectomy; (5) the inclusion of a measure of relative risk (RR), including odds ratios (ORs), hazard ratios (HRs), standardised incidence ratios (SIRs), and the corresponding 95% confidence intervals (CIs), or sufficient data to calculate them; (6) studies published in English language. The following were excluded: (1) letters to the editor, commentaries, conference abstracts only, cross-sectional studies, case reports, case series, clinical trials or narrative reviews/systematic reviews/meta-analyses; (2) studies conducted among children (< 18 years) or pregnant women; (3) studies including participants diagnosed with any type of cancer (except non-melanoma skin cancer) or with a history of cancer at baseline. Screening of the retrieved studies was independently conducted by two reviewers (F.L. and G.Mo.); a third reviewer was invited to resolve any conflicts (R.H.), using the Covidence systematic review software (Veritas Health Innovation, Melbourne, Australia). In addition, study authors and description of the study population were examined to determine any possibility of dataset overlap to maintain the independence assumption of meta-analysis [16]. For studies based on the same dataset, those with the larger sample sizes were included in the final analysis.

Data extracted from individual studies included: first author, date of publication, journal, study design, country of study, method of obtaining raw data, number of participants, follow-up time, assessment of exposure and outcome, risk estimates and 95% CIs for the association between each exposure category and upper gastrointestinal cancer, covariates and results of subgroup analyses. Risk estimates were extracted from fully-adjusted models. Data was extracted by one reviewer (F.L.) and checked by a second reviewer (R.H.).

This meta-analysis followed the Preferred Reporting Items for Systematic Reviews and Meta-Analyses (PRISMA) guidelines [17]. The completed PRISMA checklist is available in supplementary material [see Additional file 2]. The study protocol was registered on the International Prospective Register of Systematic Reviews (PROSPERO) platform (registration number: CRD42024563020). An amendment was made in October 2024 to update the date of the last literature search to September 2024.

### Study quality assessment

The Newcastle-Ottawa Scale (NOS) [18] was used to assess the quality of the included studies and evaluate their risk of bias. The NOS evaluates studies across three categories: the selection of the study groups, the comparability between groups, and the ascertainment of exposure or outcome of interest. Studies with a NOS score ≥ 6 were considered to have a low risk of bias; this cut-off was consistent with previous literature [19]. Study quality was assessed by one reviewer (F.L.) and checked by a second reviewer (R.H.). A breakdown of the NOS scores for each included study is provided in Supplementary Table 3-4 (see Additional file 1).

### Statistical analysis

Based on the assumption that upper gastrointestinal cancers are rare [2], all results are presented as RRs [20]. Studies that reported only subtype- or subsite-specific results were considered independent estimates and included in analyses for overall upper gastrointestinal cancer risk. Under the assumption that the effect of gallstones or cholecystectomy on upper gastrointestinal cancers are similar, but not identical, in different studies, random effects models were performed to estimate pooled RRs and corresponding 95% CIs [21]. Individual and pooled effect estimates were visualised using forest plots. Additionally, the prediction intervals from random effects models were estimated by the Higgins-Thompson-Spiegelhalter method [22]. These intervals indicate the range within which the effect size of a new study is likely to fall and were reported to enhance clinical interpretation of the findings [23].

Between-study heterogeneity was assessed using I^2^ and τ^2^ statistics, as estimated by the inverse variance method and the restricted maximum likelihood procedure [24], respectively. Subgroup analyses and meta-regression were conducted where feasible by sex, region (Asia, non-Asia), follow-up time (< 5 years, 5-9 years, ≥ 10 years), publication time (before 2012, or 2012 onwards, based on the publication year of a previous review [13]), study design (case-control study, cohort study), measure of outcome (HR, OR, SIR) and cancer subtype/subsite (oesophageal cancer: OAC or OSCC; gastric cancer: GCC or NCGC), to investigate the potential sources of heterogeneity. Publication bias was assessed using visual inspection of funnel plots and Egger’s tests [25, 26].

A series of sensitivity analyses were performed to ensure the robustness of the results: 1) leave-one-out meta-analyses to identify influential studies; 2) exclusion of studies below the NOS threshold to provide a summary estimate using data from only studies with a low risk of bias; 3) substituting estimates with the > 5-year follow-up estimate for the 2-to-5-year estimate from one study that reported results by follow-up time; 4) exclusion of studies where the cholecystectomy status of patients with gallstones was unclear [11, 27–29]; and 5) using multi-level random effects models to account for the inclusion of potentially correlated subtype- or subsite-specific estimates reported by the same studies.

All statistical analyses were performed using the ‘meta’, ‘metafor’ and ‘dmetar’ packages in R software (version 4.4.1).

## Results

As shown in **Figure 1**, database searches yielded 708 records. After removing 107 duplicates, 601 records underwent title and abstract screening, which identified 23 articles for full-text review. After applying the selection criteria, 11 studies were excluded [12–14, 27, 30–36], and 12 eligible studies were identified for inclusion [11, 29, 37–46]. Additionally, three studies were identified by citation tracking [28, 47, 48]. Thus, 15 studies were included in the review. Of these, 11 reported on oesophageal cancer [11, 28, 29, 37–40, 42, 43, 45, 48], and 13 reported on gastric cancer [11, 28, 29, 37–39, 41, 42, 44–48].

**Figure 1.**
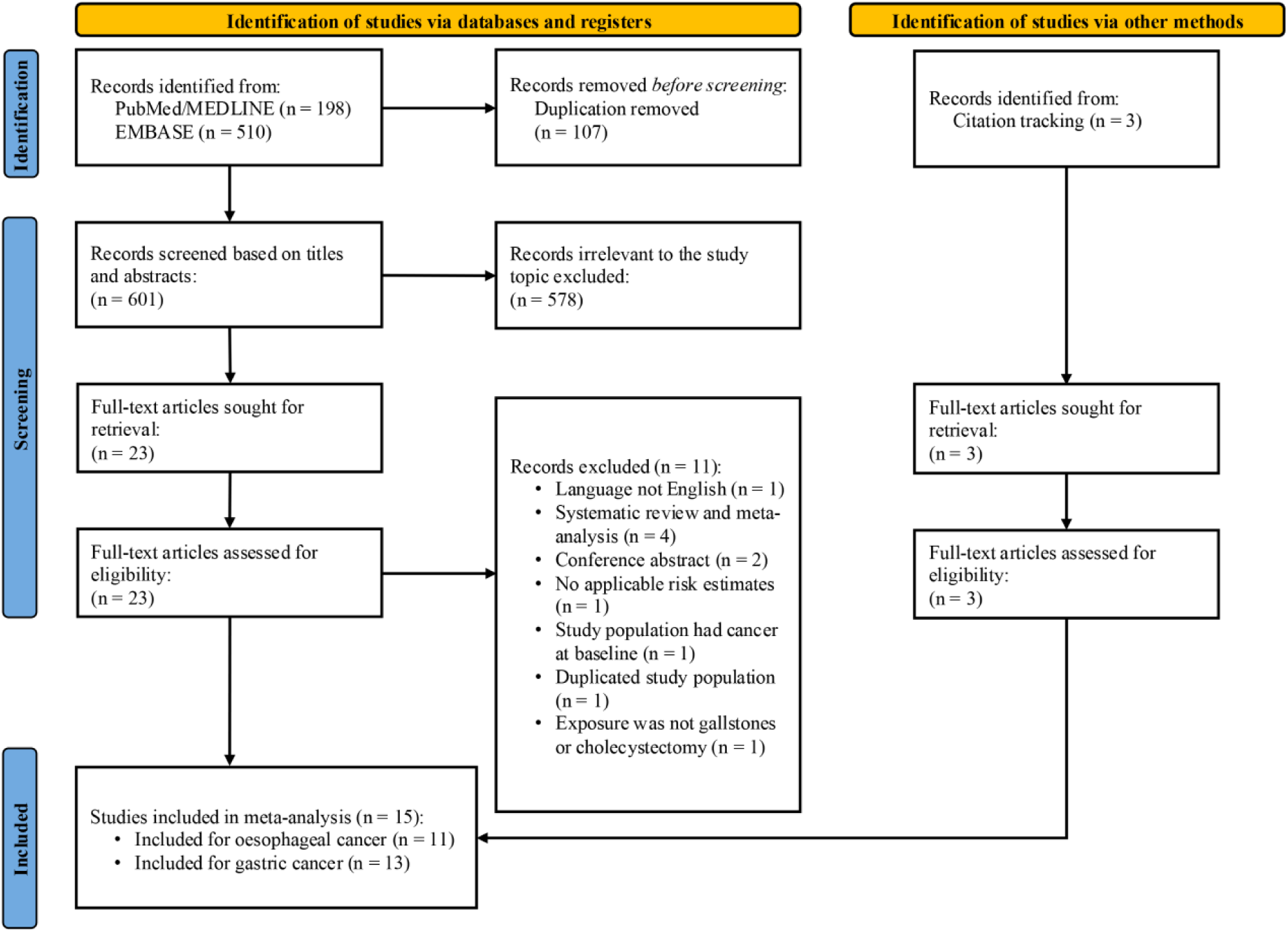
Flow diagram of study selection

### Characteristics of the included studies

Among the 11 studies included for oesophageal cancer, three were case-control studies [29, 45, 48] and eight were cohort studies. Four cohort studies reported HRs [11, 37–39], four calculated SIRs [28, 40, 42, 43], and all case-control studies reported ORs. One study had a high risk of bias [29]. The total number of oesophageal cancer cases was 12,882 among 6,036,959 participants (0.21%) (see Additional file 1: Supplementary Table 1).

Among the 13 studies which examined gastric cancer as the outcome, four were case-control studies [29, 45, 46, 48] and nine were cohort studies. Four cohort studies reported HRs [11, 37–39], five reported SIRs [28, 41, 42, 44, 47], and all case-control studies calculated ORs. Two studies had a high risk of bias [29, 47]. A total of 16,831 gastric cancer cases among 5,345,609 participants (0.31%) were included in the review (see Additional file 1: Supplementary Table 2).

For studies reporting HRs or ORs, comparator groups were those without the given exposure, whereas for those reporting SIRs the comparator was the general population.

### Associations between gallstones or cholecystectomy and oesophageal cancer

Of the 11 studies included for oesophageal cancer, nine reported on gallstones [11, 28, 29, 37–40, 43, 48], six evaluated cholecystectomy [37, 40, 42, 43, 45, 48], and three assessed a combined exposure [37, 40, 43]. No associations were observed for individual studies comparing oesophageal cancer risk among those with gallstones to those without or the general population [pooled RR = 1.05 (95% CI: 0.97 to 1.14)] with low between-study heterogeneity (I^2^ = 0%, τ^2^ = 0), and the prediction interval ranged from 0.96 to 1.15 (**Figure 2A**). Moderate evidence of publication bias was detected by Egger’s testing (*P* = 0.05) and funnel plot (see Additional file 1: Supplementary Figure 1). For cholecystectomy, one study reported an inverse association, one reported a positive association, and for the remaining studies there was no association with oesophageal cancer risk [pooled RR = 0.99 (95% CI: 0.87 to 1.13)], with moderate between-study heterogeneity (I^2^ = 60%, τ^2^ = 0.02), and the prediction interval ranged from 0.70 to 1.41 (**Figure 2B**). No indication of publication bias was found in Egger’s testing (*P* = 0.76) or funnel plot (see Additional file 1: Supplementary Figure 2). For the combined exposure, one study reported a decreased risk, one reported an increased risk, and for three there was no association with oesophageal cancer [pooled RR = 0.99 (95% CI: 0.77 to 1.28)] with substantial between-study heterogeneity (I^2^ = 78%, τ^2^ = 0.07), and the prediction interval was 0.45 to 2.20 (**Figure 2C**). Subsequent analyses were not conducted for this exposure category due to an insufficient number of available studies.

**Figure 2.**
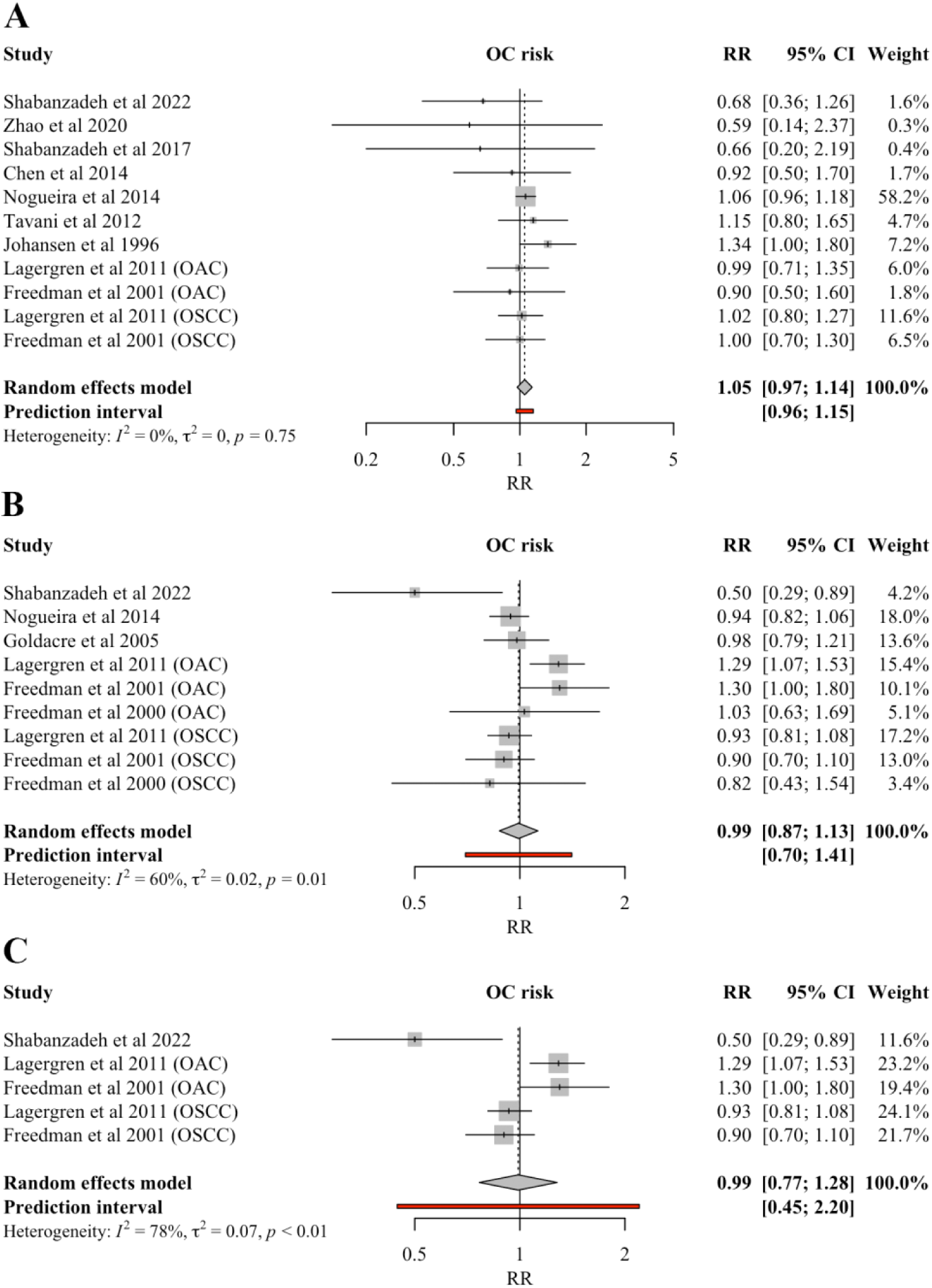
Forest plot of relative risk for oesophageal cancer across exposure categories **Panels**: (A) Gallstones; (B) Cholecystectomy; (C) Combined exposure of gallstones/cholecystectomy. **Abbreviations**: CI, confidence interval; OAC, oesophageal adenocarcinoma; OC, oesophageal cancer; OSCC, oesophageal squamous cell carcinoma; RR, relative risk.

For cholecystectomy, meta-regression analysis provided moderate evidence of variability in effect sizes by cancer subtype, with point estimates indicating a possible higher risk of OAC [pooled RR = 1.14 (95% CI: 0.94 to 1.37)] and lower risk of OSCC [pooled RR = 0.90 (95% CI: 0.81 to 1.00)], but with non-statistically significant estimates for both subgroups (*P* meta-regression = 0.02). No significant associations were found in remaining subgroup analyses for the association between gallstones or cholecystectomy and oesophageal cancer risk **(Figure 3**).

**Figure 3.**
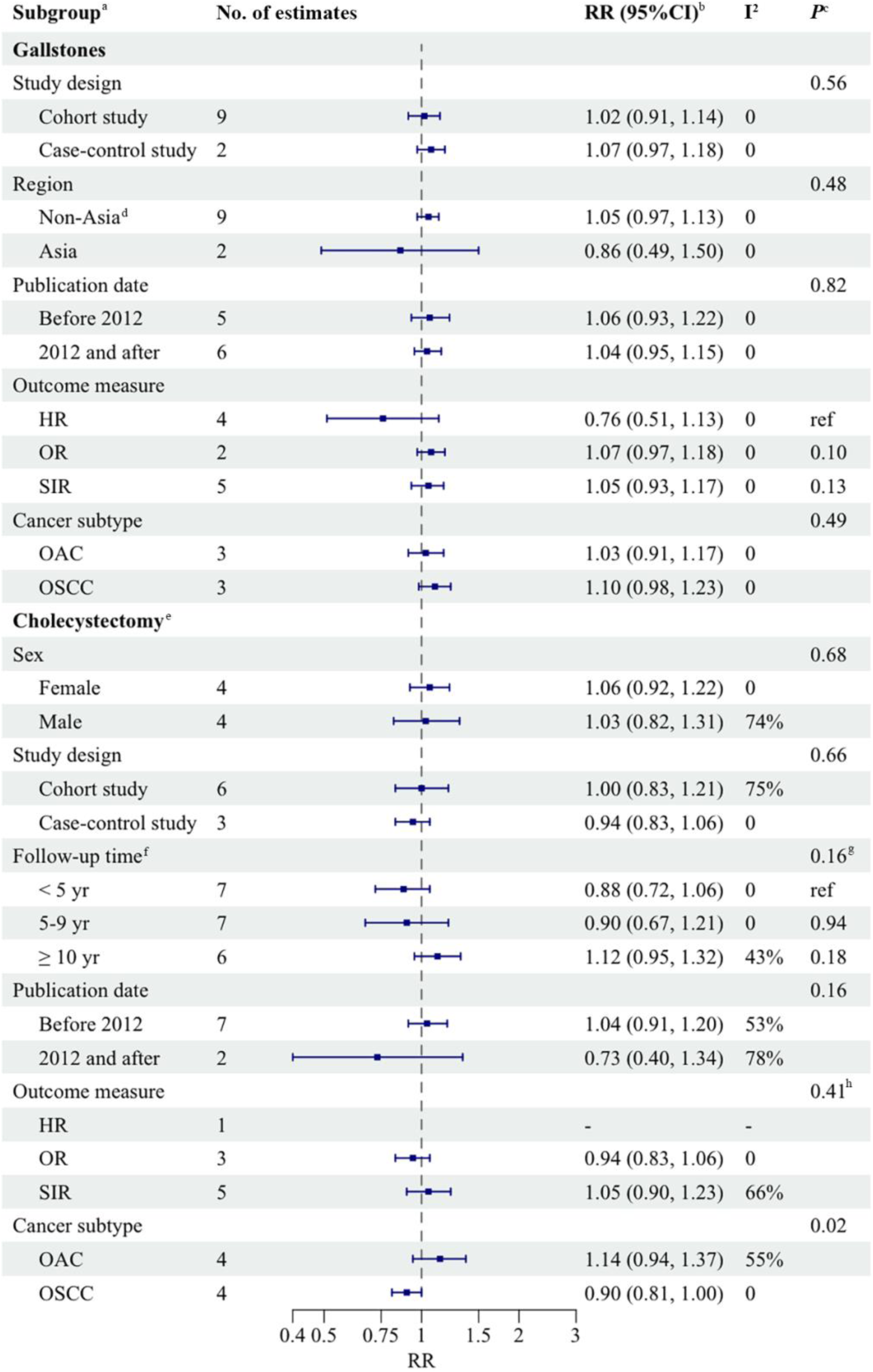
Subgroup analysis of the associations between gallstones or cholecystectomy and risk of oesophageal cancer **Abbreviations**: CI, confidence interval; HR, hazard ratio; OAC, oesophageal adenocarcinoma; OR, odds ratio; OSCC, oesophageal squamous cell carcinoma; ref, reference; RR, relative risk; SIR, standardised incidence ratio; yr, year. ^a^ Subgroups for sex and follow-up time for gallstone disease, and region for cholecystectomy were not included due to the limited number of available studies ^b^ Calculated using random effects models, with studies within each subgroup weighted by inverse variance ^c^ *P*-value for heterogeneity between subgroups in meta-regression analysis ^d^ Includes Europe and the US ^e^ Represents both cholecystectomy and combined exposure of gallstones/cholecystectomy ^f^ Categories were defined based on the most common stratification approach across included studies. For Shabanzadeh et al 2022, > 5 year follow-up estimate was grouped with 5-9 year category; for Freedman et al 2000, 5-10 year follow-up estimate was grouped with 5-9 year category ^g^ *P*-value for trend ^h^ Did not include the HR category due to the small number of studies in the subgroup

Sensitivity analyses, including leave-one-out meta-analysis, exclusion of influential studies or those below the NOS threshold, using estimates from longer follow-up period reported by Shabanzadeh *et al*. [37], exclusion of studies where the cholecystectomy status of patients with gallstones was unclear, and multi-level random effects models, did not demonstrate any considerable change in the results for all exposure categories (data not shown).

### Associations between gallstones or cholecystectomy and gastric cancer

Among the 13 studies included for gastric cancer, eight reported on gallstones [11, 28, 29, 37–39, 46, 48]; of these, three showed an increased risk, while the remaining reported no associations between gallstones and gastric cancer risk. The combined RR showed an increased risk of gastric cancer among those with gallstones [pooled RR = 1.25 (95% CI: 1.08 to 1.44)]. The prediction interval ranged from 0.87 to 1.79 and there was moderate between-study heterogeneity (I^2^ = 44%, τ^2^ = 0.02) (**Figure 4A**). Weak evidence of publication bias was shown by Egger’s testing (*P* = 0.09) and funnel plot (see Additional file 1: Supplementary Figure 3). In subgroup analyses (**Figure 5**), point summary estimates across all strata were consistently above one. Meta-regression analysis revealed significant variability in effect sizes by region, with a statistically significant positive association observed in non-Asian countries [pooled RR = 1.16 (95% CI: 1.09 to 1.23)] and weaker evidence of an association in Asian countries [pooled RR = 1.49 (95% CI: 0.95 to 2.33)] (*P* meta-regression = 0.03) (**Figure 5**).

**Figure 4.**
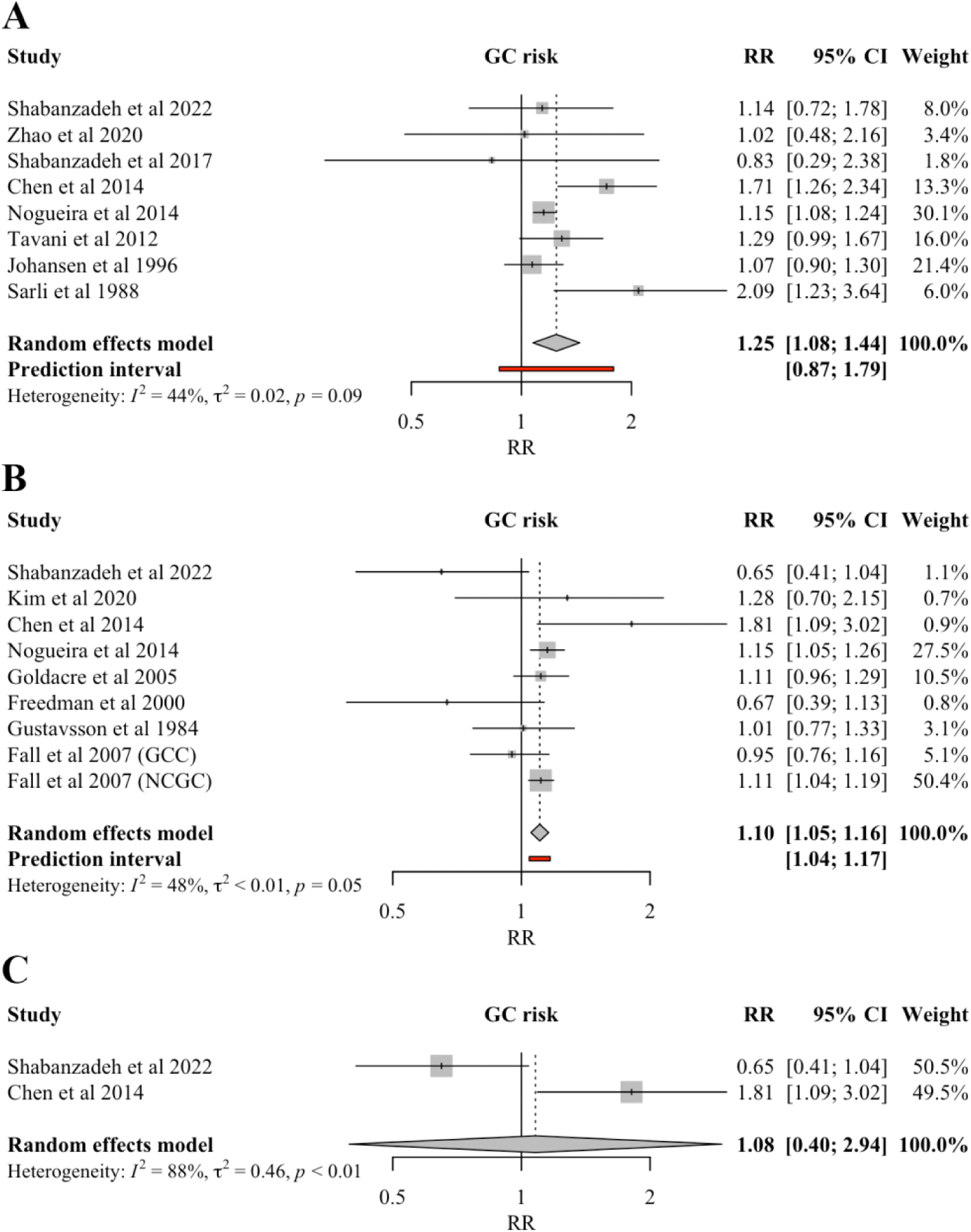
Forest plot of relative risk for gastric cancer across exposure categories **Panels**: (A) Gallstones; (B) Cholecystectomy; (C) Combined exposure of gallstones/cholecystectomy. **Abbreviations**: CI, confidence interval; GC, gastric cancer; GCC, gastric cardia cancer; NGCC, non-cardia gastric cancer; RR, relative risk.

**Figure 5.**
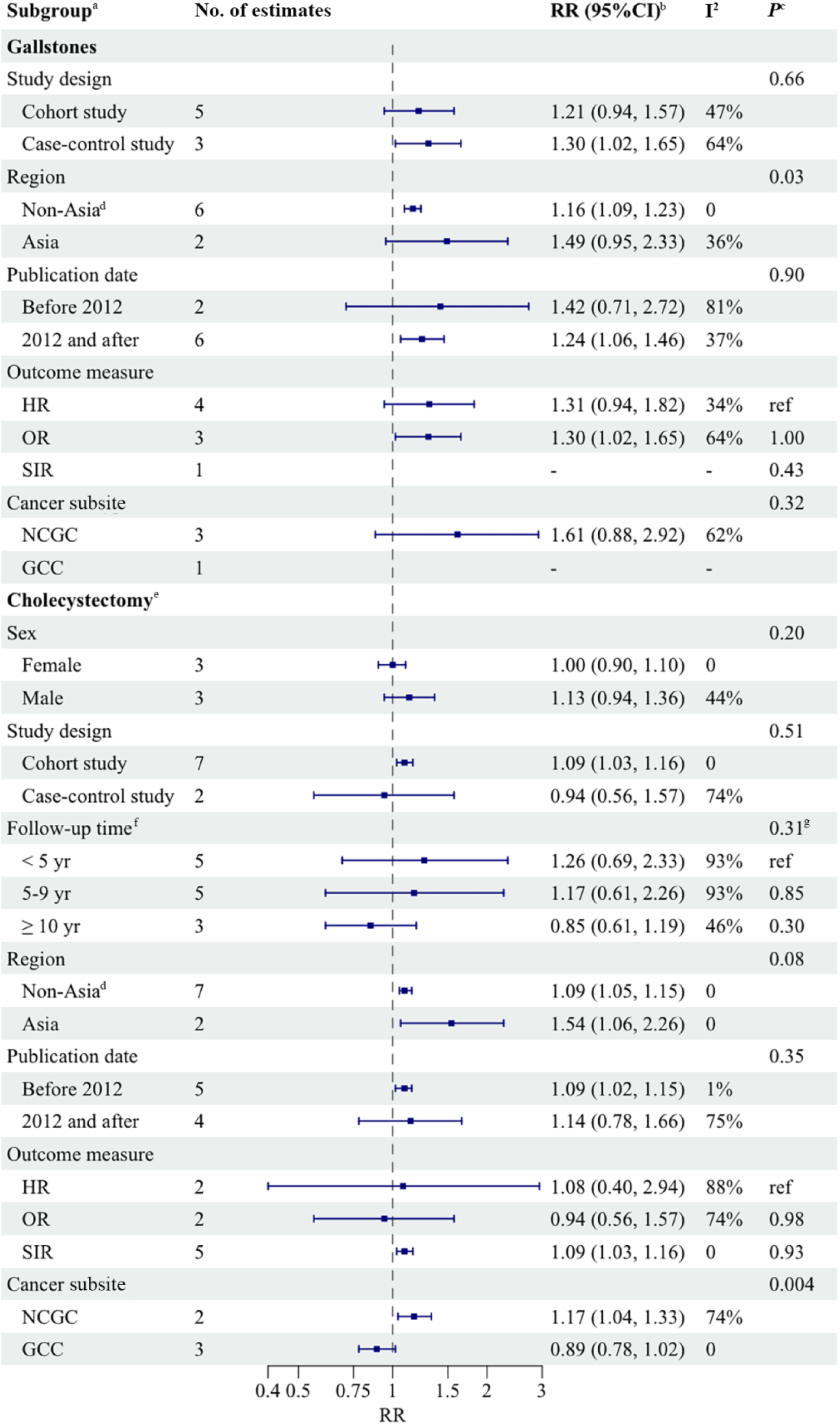
Subgroup analysis of the associations between gallstones or cholecystectomy and risk of gastric cancer **Abbreviations**: CI, confidence interval; GCC, gastric cardia cancer; HR, hazard ratio; NCGC, non-cardia gastric cancer; OR, odds ratio; ref, reference; RR, relative risk; SIR, standardised incidence ratio; yr, year. ^a^ Subgroups for sex and follow-up time for gallstone disease, and region for cholecystectomy were not included due to the limited number of available studies ^b^ Calculated using random effects models, with studies within each subgroup weighted by inverse variance ^c^ *P*-value for heterogeneity between subgroups in meta-regression analysis ^d^ Includes Europe and the US ^e^ Represents both cholecystectomy and combined exposure of gallstones/cholecystectomy ^f^ Categories were defined based on the most common stratification approach across included studies. For Shabanzadeh et al 2022 and Chen et al 2014, > 5 year follow-up estimates were grouped with 5-9 year category; for Freedman et al 2000, 5-10 year follow-up estimate was grouped with 5-9 year category ^g^ *P*-value for trend

Among the eight studies on cholecystectomy exposure [11, 37, 41, 42, 44, 45, 47, 48], three studies reported higher risks of gastric cancer, while the remainder showed no association [pooled RR = 1.10 (95% CI: 1.05 to 1.16)]. The prediction interval ranged from 1.04 to 1.17 and moderate between-study heterogeneity was detected (I^2^ = 48%, τ^2^ < 0.01) (**Figure 4B**). Evidence of publication bias was indicated by Egger’s testing (*P* = 0.04) and asymmetric funnel plot (see Additional file 1: Supplementary Figure 4). Meta-regression analysis revealed significant variability in effect sizes by cancer subsite, with a higher risk observed for NCGC [pooled RR = 1.17 (95% CI: 1.04 to 1.33)] and no association for GCC [pooled RR = 0.89 (95% CI: 0.78 to 1.02)] (*P* meta-regression= 0.004). In addition, there was moderate to weak evidence of variability in effect sizes by region, with statistically significant positive associations observed in both Asian [pooled RR = 1.54 (95% CI: 1.06 to 2.26)] and non-Asian countries [pooled RR = 1.09 (95% CI: 1.05 to 1.15)] (*P* meta-regression = 0.08) (**Figure 5**).

Two studies reported on a combined exposure of gallstones/cholecystectomy and risk of gastric cancer [11, 37], one showed a higher risk, the other showed no association. The pooled RR was 1.08 (95% CI: 0.40 to 2.94) with substantial between-study heterogeneity (I^2^ = 88%, τ^2^ = 0.46) (**Figure 3C**). Due to a limited number of studies, subsequent analyses were not performed [49].

In sensitivity analyses, using estimates from longer follow-up periods reported by Shabanzadeh *et al*. [37], the positive associations between gallstones [pooled RR = 1.19 (95% CI: 0.96 to 1.48)] or cholecystectomy [pooled RR = 1.03 (95% CI: 0.88 to 1.19)] and gastric cancer attenuated. Similarly, after excluding influential studies [41, 48], the association between cholecystectomy and gastric cancer attenuated [pooled RR = 1.01 (95% CI: 0.85 to 1.21)]. The results for other exposure categories remianed unchanged. Other sensitivity analyses, including leave-one-out meta-analysis, exclusion of studies below the NOS threshold, exclusion of studies where the cholecystectomy status of patients with gallstones was not reported, and multi-level random effects models, did not appreciably affect the risk estimates (data not shown).

## Discussion

This meta-analysis of 15 studies found that neither gallstones nor cholecystectomy were associated with oesophageal cancer risk; however, they were both associated with gastric cancer, with a 25% higher risk among those with gallstones and a 10% higher risk among those with a record of cholecystectomy compared to those without these exposures or to the general population. No associations were observed for a combined exposure of gallstones and cholecystectomy and risk of oesophageal or gastric cancer, but these analyses were based on a limited number of studies and should be interpreted with caution. Subgroup analyses suggested a possible higher risk of OAC and a lower risk of OSCC, but neither subtype-specific summary estimate reached statistical significance. We found evidence that the risk associated with cholecystectomy was higher for NCGC compared to GCC. There was also some evidence that gastric cancer risk associated with either gallstones or cholecystectomy was higher in Asian countries compared to Europe and the US.

To our knowledge, no systematic review has evaluated the effect of gallstones on oesophageal cancer, and the findings from available observational studies have been mixed. For cholecystectomy, our meta-analysis included six additional studies compared to the previously published meta-analysis, and our overall finding of no association with oesophageal cancer risk is consistent with this previous analysis published in 2012 [13]. The risk estimates reported by included studies varied significantly. Notably, Shabanzadeh *et al*. [37] reported a 50% lower risk of oesophageal cancer (of unspecified histology) associated with cholecystectomy, diverging from other studies, where the RRs ranged from 0.82 to 1.30. This might be related to the study’s unique methodological approaches, including comprehensive adjustments for a broad array of socioeconomic variables, and the use of time-varying covariates, which could allow for more accurate adjustment of confounders over time. However, mechanisms underlying this inverse association in this particular study are unclear.

Despite an observed null association between cholecystectomy and overall oesophageal cancer risk, we note with interest that the associations reported for OAC and OSCC appear to diverge, with cholecystectomy associated with a possible higher risk of OAC and lower risk of OSCC. This divergence may be explained by the different aetiology of these tumours. The possible higher risk of OAC in individuals who had undergone cholecystectomy, is consistent with a previous meta-analysis, even after the inclusion of two additional studies in our review [13]. GORD and Barrett’s oesophagus are subtype-specific risk factors for OAC [50]. A Mendelian randomisation study showed that gallstone disease is causally associated with GORD and Barrett’s oesophagus [51], indicating a potential pathway from cholecystectomy to OAC through the induction of GORD and Barrett’s oesophagus. The mechanism for the weak inverse association between cholecystectomy and OSCC remains unclear.

This is the first meta-analysis to summarise evidence on the association between gallstones and gastric cancer. Regarding exposure to cholecystectomy, this updated meta-analysis included a recently published cohort study, and the summary estimate remained consistent with two previous meta-analyses published in 2022 [12, 14]. Furthermore, in seven of the nine included studies, the exposure and outcome were ascertained through electronic medical records or registers, which likely captured the vast majority of cholecystectomy and gastric cancer cases in the study regions, enhancing their validity.

Our findings by cancer subsite aligns with prior meta-analyses [12, 14] on cholecystectomy and gastric cancer, suggesting a higher risk of NCGC after cholecystectomy. The two studies included in the NCGC stratum were large-scale studies based on Western populations with a low risk of bias, consistently showing positive associations between cholecystectomy and NCGC [41, 48]. This suggests there is value in distinguishing gastric cancer subsites in future research. More large-scale longitudinal studies and *in vivo* mechanistic studies are warranted to clarify causality of this observed association. However, it is also important to note that this finding was based on a limited number of studies and therefore should be interpreted with caution.

Disrupted bile release patterns as a result of gallstones or cholecystectomy can expose the gastric mucosa to bile acids potentially inducing oxidative stress, DNA damage, and mutations upon direct contact [52]. In line with this, we note that cancer risk decreases with increasing distance from the major duodenal papilla (where the common bile duct drains into the duodenum), with associations observed for gastric cancer but not for overall oesophageal cancer risk (but there was suggestive evidence of a higher risk of OAC, typically located in the lower thirds of the oesophagus, and a lower risk of OSCC, which usually arises in the upper two-thirds of the oesophagus [53]). We also note the distinct, though overlapping, aetiologies of gastric cancer subsites: while GCC is strongly associated with GORD (this is some evidence of associations with *Helicobacter pylori* infection, but largely limited to Asian populations [5, 54]), and NCGC with *Helicobacter pylori* infection. Notably, *Helicobacter pylori* and other *Helicobacter* species have also been shown to promote gallstone formation and biliary inflammation in mice [55], and were associated with hepatobiliary malignancies in humans [56–58]. Thus, the observed higher risk of NCGC associated with cholecystectomy, compared to GCC, may be explained by concurrent *Helicobacter* infection, potentially causing both bile dysregulation and chronic inflammation of the stomach.

We observed heterogeneity in the association between cholecystectomy and gastric cancer risk between Asian and non-Asian regions (i.e., Europe and the US), which was consistent with prior meta-analyses [12, 14]. This regional difference may be explained by multiple factors such as genetics [59], *Helicobacter pylori* prevalence [60] and strain types [61], and exposure to other risk factors including dietary habits, alcohol consumption, and smoking [54]. This finding might also be due to detection bias, with screening for gastric cancer being more rigorous in Asia compared to other parts of the world [62, 63]. However, these results should be interpreted with caution since only two studies were conducted in Asia, with one falling below the NOS threshold for a high risk of bias [47].

To our knowledge, this is the first systematic review and meta-analysis to focus on the risk of upper gastrointestinal cancers in relation to gallstones, which is one of the most prevalent digestive disorders worldwide [64]. Additionally, this study updated the prior meta-analyses on the associations between cholecystectomy and upper gastrointestinal cancers [12–14], adding large-scale studies with low risk of bias. The large sample size of this meta-analysis yielded sufficient statistical power to detect small effect sizes. Between-study heterogeneity was assessed and reported, providing a detailed exploration of potential sources of heterogeneity influencing the study findings. Finally, a thorough sensitivity analysis was performed to ensure the robustness of the results.

The results of our meta-analysis may be subject to confounding, as most included studies adjusted only for age and sex; 11 of the included studies relied on national registers and medical records, which often do not provide individual lifestyle or anthropometric data. Additional studies with comprehensive adjustment for confounders are needed to obtain more accurate effect sizes. Some studies included in this analysis may have been susceptible to selection bias. Of all included studies, one cohort study recruited participants from a single hospital [47], which may limit the representativeness of the general population; two case-control studies were hospital-based [29, 46], where the controls may be less representative of the population that gave rise to the upper gastrointestinal cancer cases. In addition, 10 of 11 included cohort studies did not report the extent of loss to follow-up, which limited our ability to assess the risk of attrition bias. Misclassification might have occurred in three studies where the exposure or outcome status was not ascertained with medical records or blinded assessments [29, 45, 47]. Despite our efforts to disentangle the effects of cholecystectomy from gallstones, not all studies separated these conditions, which limited our ability to properly define our exposure categories. Additionally, there may be some degree of misclassification in subgroup analyses by follow-up time, as the stratification approach varied across included studies, and estimates from three studies were grouped into the closest matching subgroups. Finally, the limited number of studies included in the meta-analysis constrained our ability to fully examine all proposed sources of heterogeneity and detect publication bias, especially for the analyses of combined exposure.

### Conclusions

This systematic review and meta-analysis showed that neither gallstones nor cholecystectomy were associated with overall oesophageal cancer risk (despite some limited evidence suggesting a divergence in risk by subtype), whereas they were both positively associated with gastric cancer, with evidence of a stronger association between cholecystectomy and NCGC compared to GCC risk. These findings may help risk stratification efforts and could be used to inform screening programmes to reduce incidence and mortality of gastric cancer.

## Data Availability

The data supporting this article are available from the corresponding author upon reasonable request.

## List of abbreviations

OAC: oesophageal adenocarcinoma
OSCC: oesophageal squamous cell carcinoma
GCC: gastric cardia cancer
NCGC: non-cardia gastric cancer
GORD: gastro-oesophageal reflux disease
RR: risk ratio
OR: odds ratio
HR: hazard ratio
SIR: standardised incidence ratio
CI: confidence interval
PRISMA: Preferred Reporting Items for Systematic reviews and Meta-Analyses
PROSPERO: International Prospective Register of Systematic Reviews
NOS: Newcastle-Ottawa Scale

## Declarations

## Acknowledgements

The authors would like to thank the librarians at Imperial College London for their assistance with literature search strategy development and access to articles.

## Authors’ contributions

F.L.: conceptualisation, methodology, investigation, formal analysis, visualisation, writing – original draft, writing – review & editing. G.Mo.: writing – review & editing. G.Mu.: investigation, writing – review & editing. A.C.: conceptualisation, writing – review & editing, supervision. R.H.: conceptualisation, investigation, writing – review & editing, supervision. All authors read and approved the final manuscript.

## Ethics approval and consent to participate

Not applicable.

## Consent for publication

Not applicable.

## Competing interests

The authors declare that they have no competing interests.

## Funding

This research was supported by Cancer Research UK (Renewing Programme Award: PRCRPG-Nov22/100001). Infrastructure support for this work was provided by the NIHR Imperial Biomedical Research Centre. The funders had no role in the study design, data collection, analysis, or interpretation, manuscript writing, or decision to submit for publication. The views expressed are those of the authors and not necessarily those of the Department of Health, NIHR, or Cancer Research UK.

## Notes

### Competing Interest Statement

The authors have declared no competing interest.

### Author Declarations

Source data were publicly available before the initiation of the study and were identified through systematic searches of PubMed and EMBASE. All data used in the meta-analysis were extracted from the published articles included in the systematic review, which are accessible via the respective journals.

