## Additional file 1 for "Association between Gallstones or Cholecystectomy and Upper Gastrointestinal Cancers: A Systematic Review and Meta-analysis"

Supplementary material

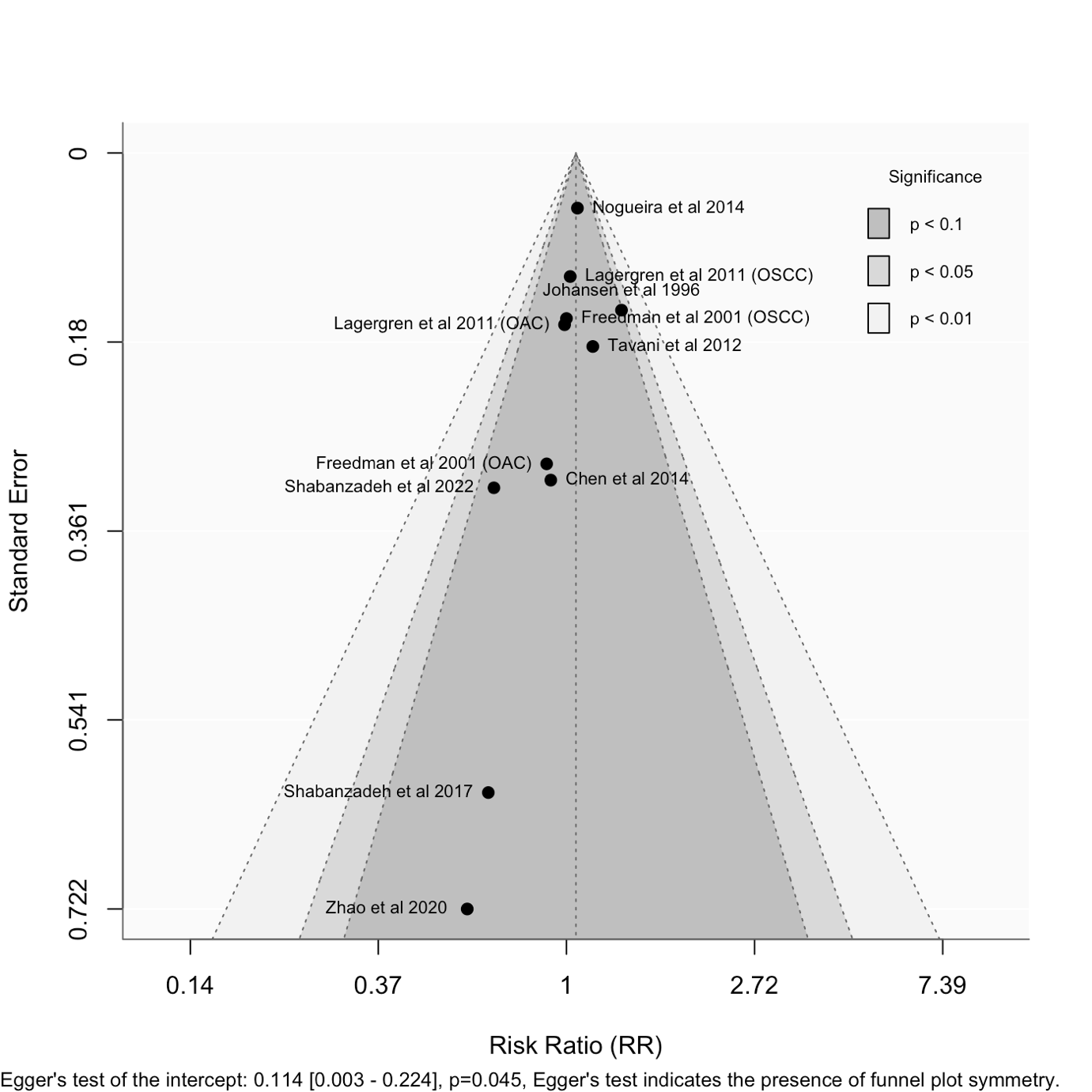

**Supplementary Figure 1**. Funnel plot for analyses of gallstones and oesophageal cancer risk.

**Abbreviations**: OAC, oesophageal adenocarcinoma; OSCC, oesophageal squamous cell carcinoma.

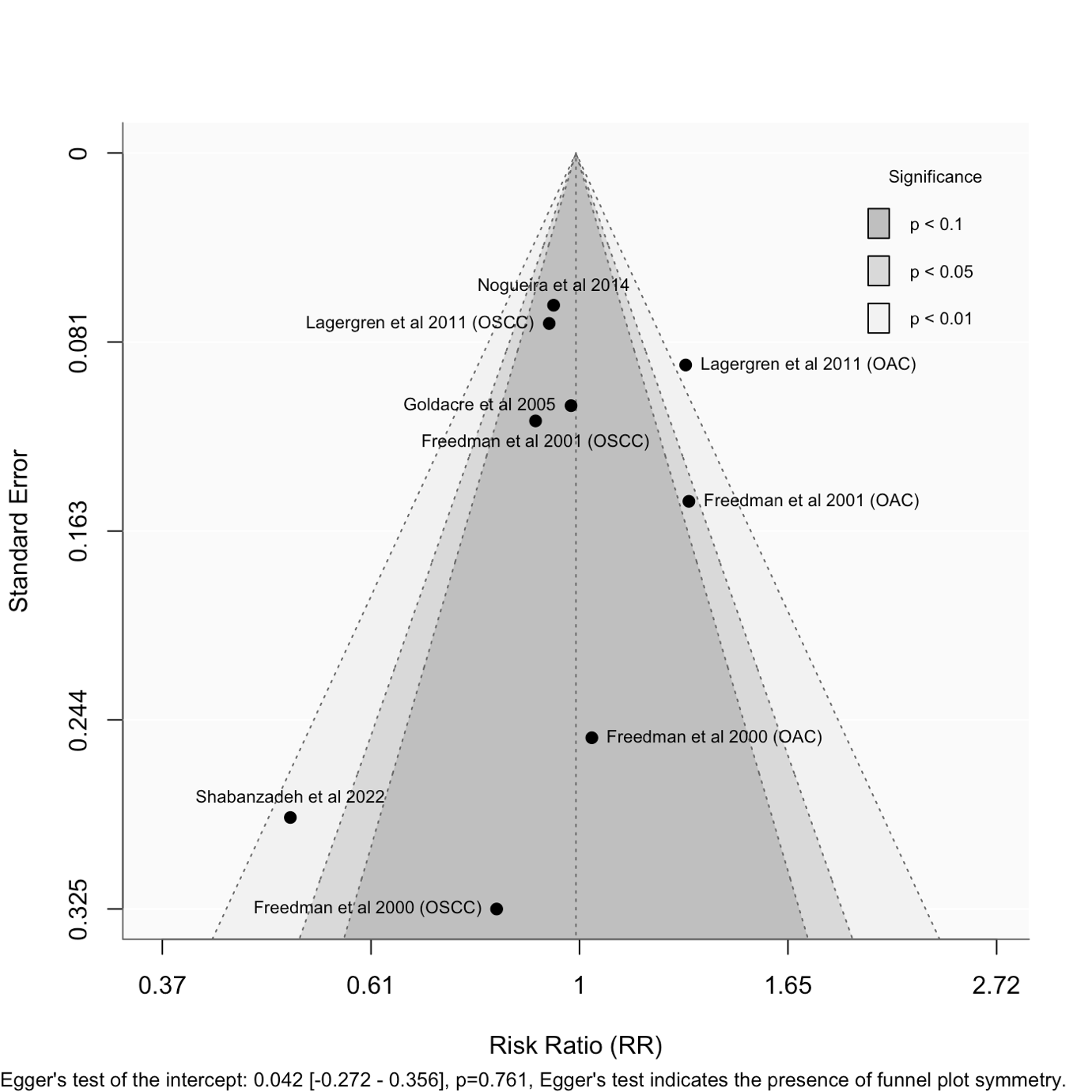

**Supplementary Figure 2**. Funnel plot for analyses of cholecystectomy and oesophageal cancer risk.

**Abbreviations**: OAC, oesophageal adenocarcinoma; OSCC, oesophageal squamous cell carcinoma.

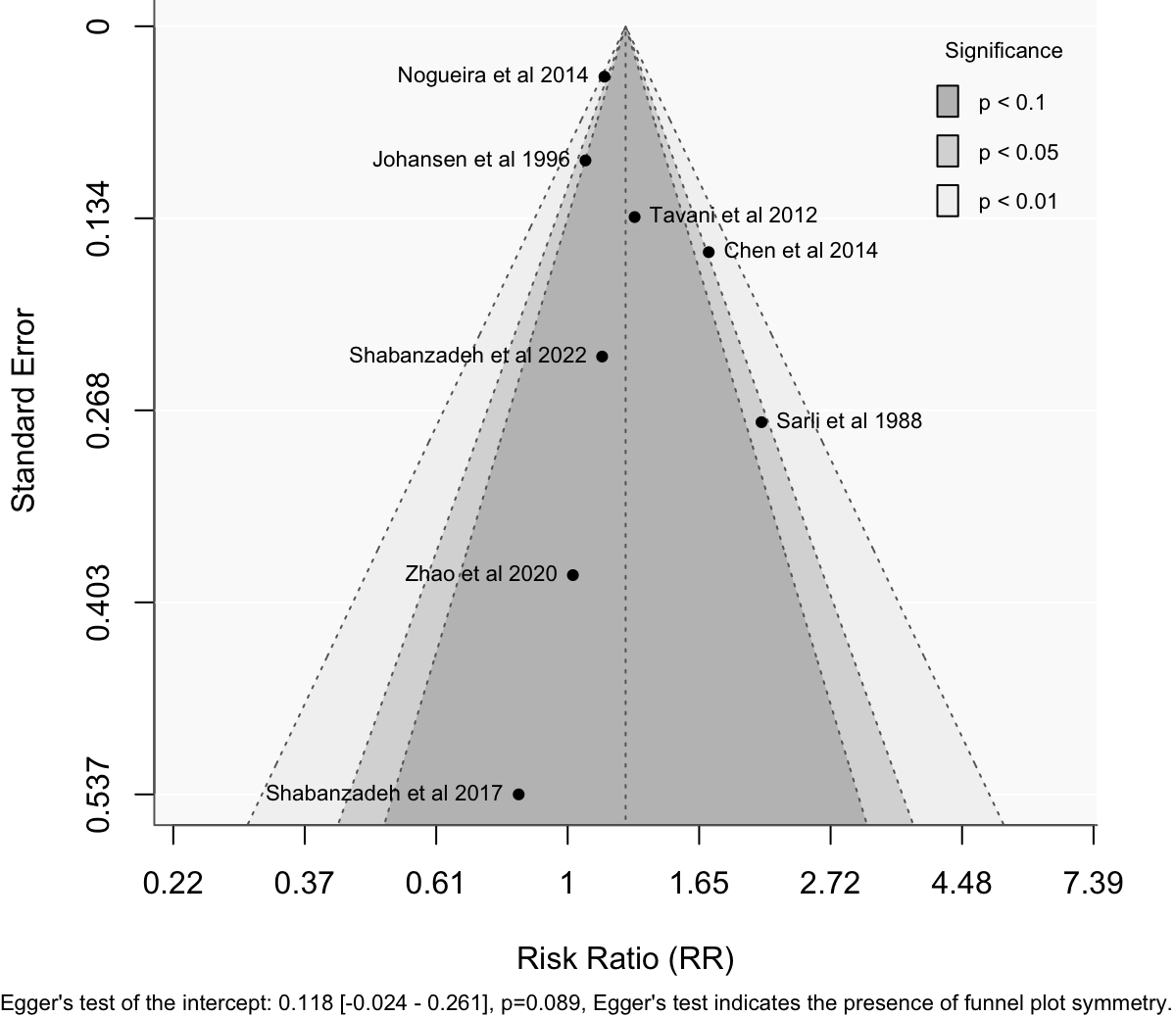

**Supplementary Figure 3**. Funnel plot for analyses of gallstones and gastric cancer risk.

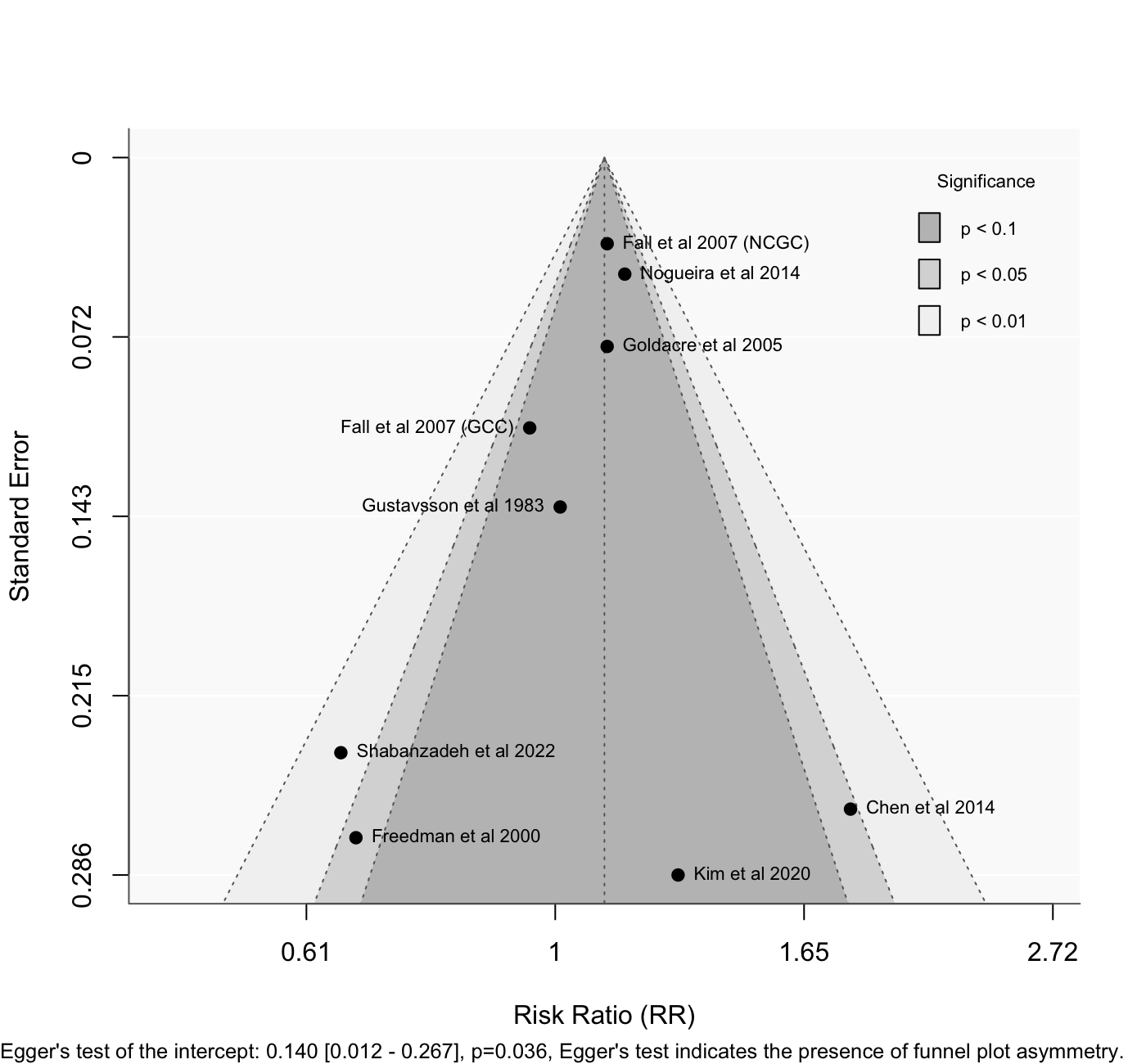

**Supplementary Figure 4**. Funnel plot for analyses of cholecystectomy and gastric cancer risk.

**Abbreviations**: NCGC, non-cardia gastric cancer; GCC, gastric cardia cancer.

| **Supplementary Table 1.** Characteristics of studies included in meta-analyses for oesophageal cancer, ordered by study design and then publication year | | | | | | | | | |
| --- | --- | --- | --- | --- | --- | --- | --- | --- | --- |
| **Author, year, country** | **Study period** | **Exposure** | **Data source** | **Number of oesophageal cancer cases** | **Size of study population^b^** | **Follow-up time** | **Outcome measure** | **Covariates** | **NOS score** |
| *Cohort studies* | | | | | | | | | |
| Shabanzadeh et al, 2022, Denmark (37) | 1977 to 2014 | GD, CE, combined^a^ | National Patient Registry and National Register of Causes of Death | 9,205 | 4,465,962 | 1977 to 2014 | HR | Sex, socioeconomic status, civil status, level of education, personal annual income | 8 |
| Zhao et al, 2020, China (38) | 2006 to 2007 | GD | Data obtained via medical examination, questionnaire and linked medical insurance system | 135 | 79, 809 | 11 years (median) | HR | Education, smoking status, drinking status, diabetes, fatty liver levels, BMI | 9 |
| Shabanzadeh et al, 2017, Denmark (39) | 1982 to 2014 | GD | Data obtained via medical examination, questionnaire, National Patient registry and National Register of Causes of Death | 34 | 5,928 | 24.7 years (median) | HR | Age | 6 |
| Chen et al, 2014, Taiwan, China (11) | 2000 to 2010 | GD | National health insurance database | 65 | 77,725 | >5 years | HR | Age, sex, number of comorbidities | 8 |
| Lagergren et al, 2011, Sweden (40) | 1965 to 2008 | GD, CE, combined^a^ | National Inpatient and Cancer Registers | CE cohort: 126 OAC cases, 193 OSCC cases; Unoperated cohort with gallstones: Not reported | CE cohort: 345,251; Unoperated cohort with gallstones: 192,960 | CE cohort: 15 years (mean);  Unoperated cohort with gallstones: 9 years (mean) | SIR | Age, sex, calendar year | 8 |
| Goldacre et al, 2005, UK (42) | 1963 to 1999 | CE | Hospital records | 894 | 374,067 | ≥10 years | SIR | Age, sex, calendar year of first recorded admission, district of residence | 8 |
| Freedman et al, 2001, Sweden (43) | 1965 to 1997 | GD, CE, combined^a^ | National Inpatient and Cancer Registers | CE cohort: 53 OAC cases, 129 OSCC cases; Cholelithiasis cohort: Not reported | CE cohort: 268,312; Cholelithiasis cohort: 167,646 | CE cohort: 13 years (mean);  Cholelithiasis cohort: 5 years (mean) | SIR | Age, sex, calendar year | 8 |
| Johansen et al, 1996, Demark (28) | 1977 to 1989 | GD | Hospital Discharge Register and National Cancer Registry | 41 | 42,098 | 7.4 years (mean) | SIR | Age, sex, calendar year | 7 |
| *Case-control studies* | | | | | | | | | |
| Nogueira et al, 2014, US (48) | 1992 to 2005 | GD, CE | Cases identified via linked cancer registries to health insurance data; controls identified via health insurance data | Not reported | 11,442^c^ | NA | OR | Age, sex, calendar year of selection, duration of Medicare benefits coverage | 9 |
| Tavani et al, 2012, Italy and Switzerland (29) | 1982 to 2009 | GD | Hospital records, questionnaire, and self-reporting | 917 cases | 3,666 controls | NA | OR | Sex, quinquennia of age, study centre, year of interview, study period, education, alcohol drinking, tobacco smoking, BMI | 4 |
| Freedman et al, 2000, Sweden (45) | 1995 to 1997 | CE | Cases identified via hospital records and regional tumour registries; controls identified via national population register | 189 OAC cases, 167 OSCC cases | 820 controls | NA | OR | Age, sex, tobacco use, alcohol use, BMI, educational level, intake of fruit and vegetables, meal size, physical activity | 7 |
| **Abbreviations**: BMI, body mass index; CE, cholecystectomy; GD, gallstone disease; HR, hazard ratio; NOS, Newcastle-Ottawa Scale; OAC, oesophageal adenocarcinoma; OR, odds ratio; OSCC, oesophageal squamous cell carcinoma; SIR, standardised incidence ratio.  ^a^ Combined exposure of gallstones and cholecystectomy  ^b^ Presents the total size of the study population for cohort studies and the number of control participants for case-control studies  ^c^ Total number of participants | | | | | | | | | |

| **Supplementary Table 2.** Characteristics of studies included in meta-analyses for gastric cancer, ordered by study design and then publication year | | | | | | | | | |
| --- | --- | --- | --- | --- | --- | --- | --- | --- | --- |
| **Author, year, country** | **Study period** | **Exposure** | **Data source** | **Number of gastric cancer cases** | **Size of study population^b^** | **Follow-up time** | **Outcome measure** | **Covariates** | **NOS score** |
| *Cohort studies* | | | | | | | | | |
| Shabanzadeh et al, 2022, Denmark (37) | 1960 to 2014 | GD, CE, combined^a^ | National Patient Registry and National Register of Causes of Death | 10,371 | 4,465,962 | 1977 to 2014 | HR | Sex, socioeconomic status, civil status, level of education, personal annual income | 8 |
| Zhao et al, 2020, China (38) | 2006 to 2007 | GD | Data obtained via medical examination, questionnaire and linked medical insurance system | 239 | 79, 809 | 11 years (median) | HR | Education, smoking status, drinking status, diabetes, fatty liver levels, BMI, site for physical examination, physical activity, total cholesterol, high-density lipoprotein cholesterol (HDL-C), triglyceride | 9 |
| Kim et al, 2020, Korea (47) | 2006 to 2013 | CE | Hospital records | 14 | 3,588 | 15 months (median) | SIR | Not reported | 3 |
| Shabanzadeh et al, 2017, Denmark (39) | 1982 to 2014 | GD | Data obtained via medical examination, questionnaire, National Patient registry and National Register of Causes of Death | 38 | 5,928 | 24.7 years (median) | HR | Age | 6 |
| Chen et al, 2014, Taiwan, China (11) | 2000 to 2010 | GD, CE, combined^a^ | National health insurance database | GD: 202 cases, CE and combined: 60 cases | 77,725 | >5 years | HR | Age, sex, number of comorbidities | 8 |
| Fall et al, 2007, Sweden (41) | 1970 to 1997 | CE | National Inpatient and Cancer Registers | 948 | 251,672 | 11.5 years (mean) | SIR | Age, sex, calendar year | 8 |
| Goldacre et al, 2005, UK (42) | 1963 to 1999 | CE | Hospital records | 1,531 | 374,067 | ≥10 years | SIR | Age, sex, calendar year of first recorded admission, district of residence | 8 |
| Johansen et al, 1996, Demark (28) | 1977 to 1989 | GD | Hospital Discharge Register and National Cancer Registry | 118 | 42,098 | 7.4 years (mean) | SIR | Age, sex, calendar year | 7 |
| Gustavsson et al, 1984, Sweden (44) | 1964 to 1967 | CE | National Inpatient and Cancer Registers | 89 | 16,773 | Range: 11 to 14 years | SIR | Age, sex | 7 |
| *Case-control studies* | | | | | | | | | |
| Nogueira et al, 2014, US (48) | 1992 to 2005 | GD, CE | Cases identified via linked cancer registries to health insurance data; controls identified via health insurance data | Not reported | 22,860^c^ | NA | OR | Age, sex, calendar year of selection, duration of Medicare benefits coverage | 9 |
| Tavani et al, 2012, Italy and Switzerland (29) | 1982 to 2009 | GD | Hospital records, questionnaire, and self-reporting | 999 cases | 2,628 controls | NA | OR | Sex, quinquennia of age, study centre, year of interview, study period, education, alcohol drinking, tobacco smoking, BMI | 4 |
| Freedman et al, 2000, Sweden (45) | 1995 to 1997 | CE | Cases identified via hospital records and regional tumour registries; controls identified via national population register | 262 cases | 820 controls | NA | OR | Age, sex, tobacco use, alcohol use, BMI, educational level, intake of fruit and vegetables, meal size, physical activity | 7 |
| Sarli et al, 1988, Italy (46) | 1982 to 1985 | GD | Hospital records | 209 cases | 209 controls | NA | OR | Age, sex, geographical region, dietary habit | 6 |
| **Abbreviations:** BMI, body mass index; CE, cholecystectomy; GD, gallstone disease; HDL-C, high-density lipoprotein cholesterol; HR, hazard ratio; OR, odds ratio; SIR, standardised incidence ratio.  ^a^ Combined exposure of gallstones and cholecystectomy  ^b^ Presents the total size of the study population for cohort studies and the number of control participants for case-control studies.  ^c^ Total number of participants | | | | | | | | | |

| **Supplementary Table 3.** Breakdown of Newcastle-Ottawa scale score for included cohort studies. | | | | | | | | | | | |
| --- | --- | --- | --- | --- | --- | --- | --- | --- | --- | --- | --- |
| **Study** | **Cancer type** | **Selection** | | | | **Comparability**^a^ | **Outcome** | | | **Total Score** | **Risk of bias** |
|  |  | Q1 | Q2 | Q3 | Q4 |  | Q1 | Q2^b^ | Q3^c^ |  |  |
| Shabanzadeh et al 2022 (37) | OC & GC | * | * | * | * | ** | * | * | - | 8 | Low |
| Zhao et al 2020 (38) | OC & GC | * | * | * | * | ** | * | * | * | 9 | Low |
| Kim et al 2020 (47) | GC | - | - | * | * | - | - | * | - | 3 | High |
| Shabanzadeh et al 2017 (39) | OC & GC | * | * | * | * | - | * | * | - | 6 | Low |
| Chen et al 2014 (11) | OC & GC | * | * | * | * | ** | * | * | - | 8 | Low |
| Lagergren et al 2011 (40) | OC | * | * | * | * | ** | * | * | - | 8 | Low |
| Fall et al 2007 (41) | GC | * | * | * | * | ** | * | * | - | 8 | Low |
| Goldacre et al 2005 (42) | OC & GC | * | * | * | * | ** | * | * | - | 8 | Low |
| Freedman et al 2001 (43) | OC | * | * | * | * | ** | * | * | - | 8 | Low |
| Johansen et al 1996 (28) | OC & GC | * | * | * | * | * | * | * | - | 7 | Low |
| Gustavsson et al 1984 (44) | GC | * | * | * | * | * | * | * | - | 7 | Low |
| **Abbreviations**: GC, gastric cancer; OC, oesophageal cancer.  ^a^ One star was awarded if the study controlled for both age and sex. A second star was awarded if any additional factor was accounted for  ^b^ A minimum follow-up duration of five years was considered adequate for outcome occurrence based on the stratification approach used in the included studies  ^c^ A follow-up rate of 80% or higher was considered adequate | | | | | | | | | | | |

| **Supplementary Table 4.** Breakdown of Newcastle-Ottawa scale score for included case-control studies. | | | | | | | | | | | |
| --- | --- | --- | --- | --- | --- | --- | --- | --- | --- | --- | --- |
| **Study** | **Cancer type** | **Selection** | | | | **Comparability**^a^ | **Exposure** | | | **Total score** | **Risk of bias** |
|  |  | Q1 | Q2 | Q3 | Q4 |  | Q1 | Q2 | Q3 |  |  |
| Nogueira et al 2014 (48) | OC & GC | * | * | * | * | ** | * | * | * | 9 | Low |
| Tavani et al 2012 (29) | OC & GC | * | - | - | - | ** | - | * | - | 4 | High |
| Freedman et al 2000 (45) | OC & GC | * | * | * | - | ** | - | * | * | 7 | Low |
| Sarli et al 1988 (46) | GC | * | - | - | * | ** | * | - | * | 6 | Low |
| **Abbreviations**: GC, gastric cancer; OC, oesophageal cancer.  ^a^ One star was awarded if the study controlled for both age and sex. A second star was awarded if any additional factor was accounted for | | | | | | | | | | | |
